# Participating in vaccine research for COVID-19 in Senegal: trust and information

**DOI:** 10.1101/2021.04.07.21255079

**Authors:** Valéry Ridde, Mouhamadou Faly Ba, Ibrahima Gaye, Amadou Ibra Diallo, Emmanuel Bonnet, Adama Faye

**Author notes:** Corresponding author. E-mail address (V. Ridde).

## Abstract

This research aims to understand the level and determinants of people’s willingness to participate in a vaccine trial for COVID-19 in Senegal. We conducted a telephone survey among a marginal quota sample of 607 people over 18 years of age. Only 44.3% of the participants wanted to participate in a vaccine trial for COVID-19, with females intending to participate more than males. Participants who intended to be vaccinated against COVID-19 (OR = 6.48, 95% CI [4.12-10.4]) and who thought that being infected with the coronavirus would have a significant impact on their health (OR = 2.34, 95% CI [1.57, 3.51]) were more likely to agree to take part in the COVID-19 vaccine research. Confidence in the vaccine, health personnel, and government in the fight against the pandemic are key factors in intending to participate in vaccine research in Senegal.

## 1. Introduction

The arrival of the COVID-19 pandemic has not changed Africa’s collective imagination and discourse on Africa as a medical and vaccine testing ground. Indeed, historians and other social scientists have long documented how Africa has been the site of biomedical research, whether for testing drugs or vaccines [1-5]. Social scientists have shown the need for a complex and nuanced view beyond the ethical challenges and power issues [2,4]. However, we are in the process of “*globalizing human subjects research*” [6] after the scientific imperialism of the Pasteur Institutes [7].

In francophone Africa, particularly in Senegal, the COVID-19 pandemic has opened a new window of opportunity for these debates. Contrary to what is imagined about infectious diseases, after Ebola [8,9] or Lassa [10], Africa was neither the first continent to be affected nor the one that, thus far, suffered the most consequences [11]. However, a televised interview with two French doctors set off the “*April Fool’s prank*” on April 1^st^, 2020 [12]. In a context where debates on the decolonisation of global health are numerous [13,14], Africans hear these two Frenchmen say that it is necessary to test the BCG vaccine for prevention in Africa against COVID-19. Academic reactions on the ethical issues this raises and reactions in Africa’s (social) media will be numerous [12,15]. Rumours about Africa as a vaccine testing ground will, from then on, continue to grow significantly [16]. At the end of the 1970s in Senegal, the first vaccine trials against hepatitis B gave rise to numerous ethical debates [3]. The 2007 meningitis vaccine trial also gave rise to numerous controversies, showing the lack of journalist’s scientific culture and an attempt at political instrumentalisation [17]. This story was brought back to the forefront by the national media in April 2020 with the French doctors’ speech. Yet Africa is by far one of the continents where biomedical research is least carried out, with COVID-19 being no exception [18]. A May 2020 analysis shows that of the 1002 therapeutic clinical trials for COVID-19 worldwide, only 32 (3.2%) were conducted in Africa [19]. Of the hundreds of ongoing vaccine trials against COVID-19, only 16 are taking place in Africa as of mid-March 2021 (South Africa: 11; Kenya = 3; Egypt 1; Morocco 1^1^).

However, like other continents [20], Africa needs to conduct research and trials to test drugs and vaccines against COVID-19 [21]. Not only is this essential to adapt biomedical products and vaccines to national contexts and populations, but countries now have research centres and ethics committees competent to carry out these trials under good ethical conditions [18]. Thus, in a context where vaccines are the common solution to fight against the COVID-19 pandemic and other epidemics to come, it is important to understand the willingness of Senegalese to participate in a vaccine trial. Qualitative research in Senegal has shown that Senegalese people do not wish to be “exploited” by such COVID-19 vaccine trials but noted an ambiguous relationship and significant ignorance about how biomedical research works [22]. Thus, the objective of this article is to understand the level and determinants of Senegal’s population’s willingness to participate in a COVID-19 vaccine trial.

## 2. Method

We conducted a cross-sectional, descriptive, and analytical study. We collected data from December 24, 2020, to January 16, 2021. We carried out the study on a sample of 607 people aged over 18 years. We used the marginal quota sampling strategy [23] to have a representative sample of the national population stratified by population weight by region, gender, and age group. Five female interviewers speaking six languages (French, Diola, Wolof, Sérére, Pulaar, Soninké) carried out data collection through a telephone survey. We used tablets equipped with Open Data Kit (ODK) software to administer the questionnaire.

The variables in our study were based on acceptability models [24,25]. They included socio-demographic characteristics (age, gender, education, region of residence, wealth tercile, chronic medical conditions), vaccination history, attitude towards the COVID-19 vaccine measured by five items in the form of a five-point Likert scale, intention to be vaccinated against COVID-19, fear of the coronavirus, concern for serious health consequences if infected by the coronavirus, trust in the government to fight the COVID-19 epidemic, willingness to participate in COVID-19 vaccine research, and reasons for refusing or agreeing to participate in this research.

We used R software version 4.0.3 for data analysis. We described the quantitative variables by the mean ± standard deviation and the qualitative variables by their frequencies. Then, we used a Ch2 test to compare two qualitative variables and a student’s t test a qualitative and a quantitative variable. Finally, we used multivariate logistic regression to determine the factors associated with the willingness of respondents to participate in COVID-19 vaccine research. All variables with p-values less than 0.25 in the comparisons were retained for the full model construction [26]. We used the stepwise top-down selection procedure to build a more parsimonious reduced model [27]. Significance was considered at a p-value < 0.05.

## 3. Results

Women represented 39.7% of the respondents. In our study, 15.5% of the respondents reported having a chronic disease. The average confidence in the Senegalese government in the fight against the epidemic was 6.6/10 ± 3.1. 54.4% of respondents expressed the intention to be vaccinated. According to their statements, 44.3% of the participants would like to participate in vaccine research for COVID-19 if it took place (Appendix 1). Of these, 23.0% (62/269) explained this because they trusted the health workers (Figure 1). Of those who refused to participate in the research, 34.4% (116/337) expressed a lack of trust in vaccines, and 29.4% (99/337) perceived a lack of safety of the vaccine research (Figure 2).

**Figure 1:**
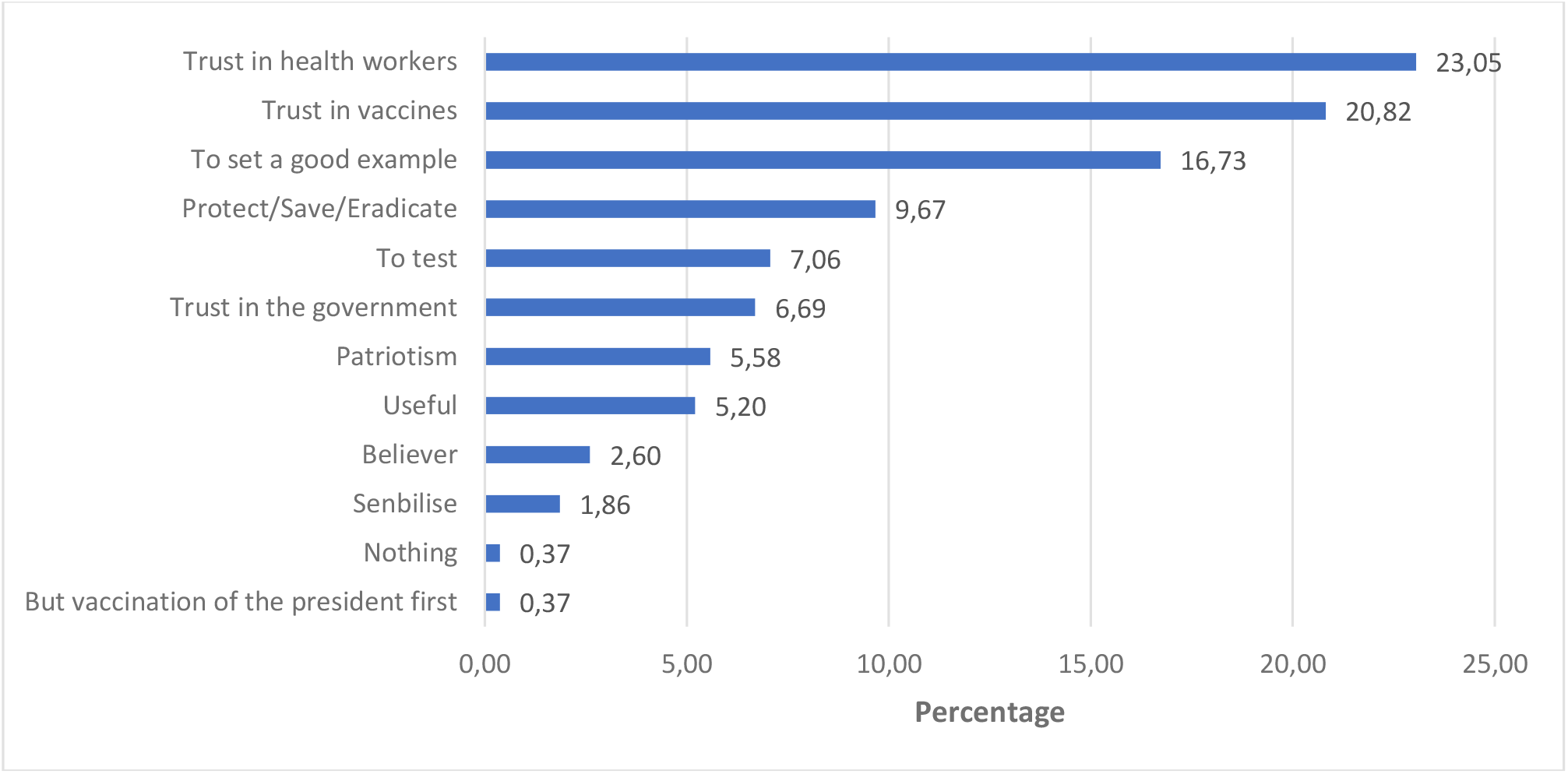
Reasons for willingness to participate in COVID-19 vaccine research (N=269)

**Figure 2:**
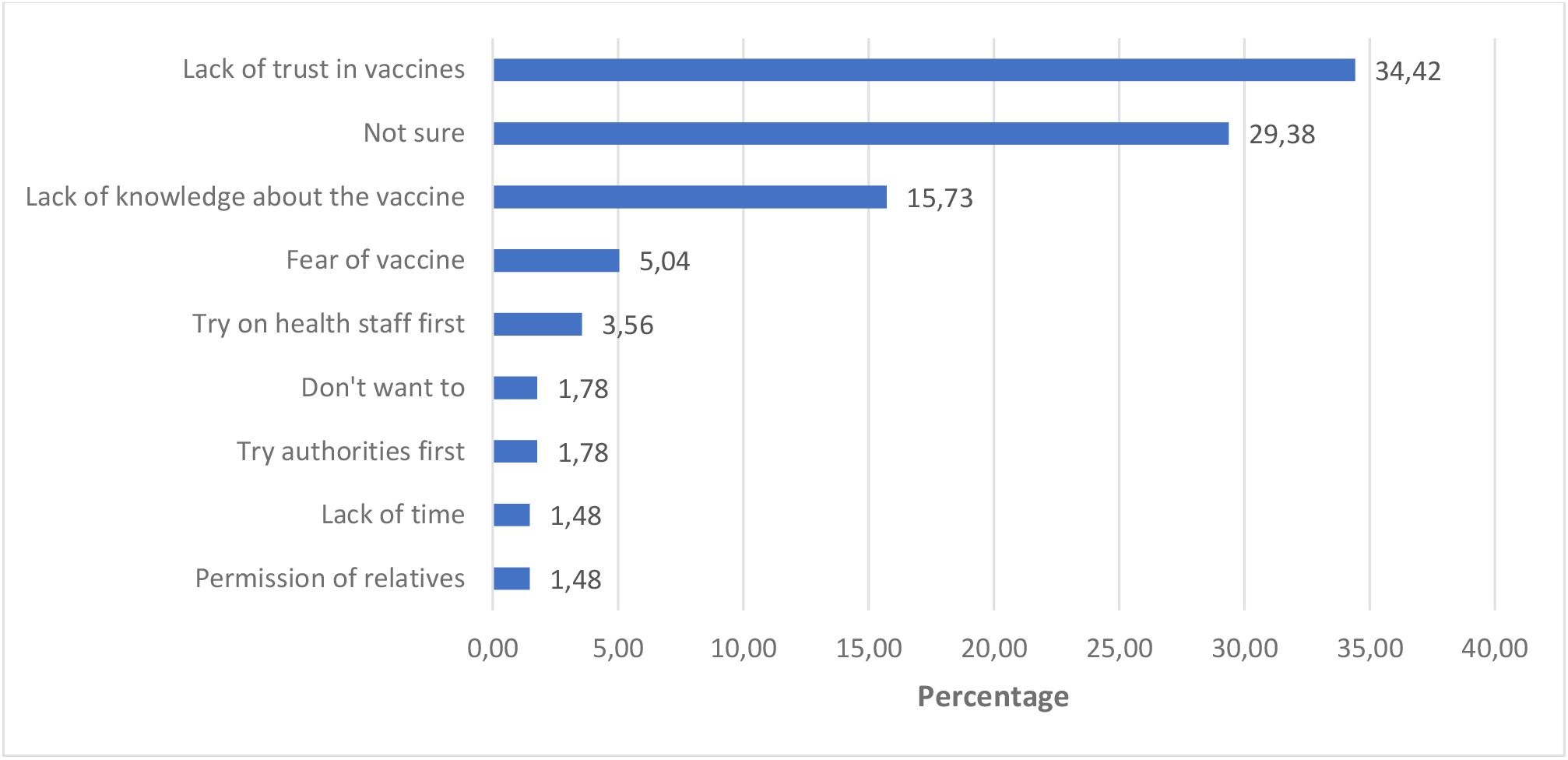
Reasons for refusing to participate in COVID-19 vaccine research (N=337)

The proportion of the men who agreed to participate in the COVID-19 vaccination research (38.5%) was significantly lower than that of the women (53.1%, p = 0.001) (Appendix 2).

The results of the multivariate analysis showed that participants who intended to be vaccinated against COVID-19 (OR = 6.48, 95% CI [4.12-10.4]) and who thought that being infected with the coronavirus would have a significant impact on their health (OR = 2.34, 95% CI [1.57, 3.51]) were more likely to agree to participate in COVID-19 vaccination research (Table 1). The other three factors positively associated with willingness to participate in COVID-19 vaccination research were being female (OR = 1.82, 95% CI [1.22-2.72]), having a positive attitude towards the vaccine (OR = 1.69, 95% CI [1.09, 2.62]), and having confidence in the Senegalese government to control the coronavirus epidemic (OR = 1.09, 95% CI [1.02, 1.16]).

**Table 1:**
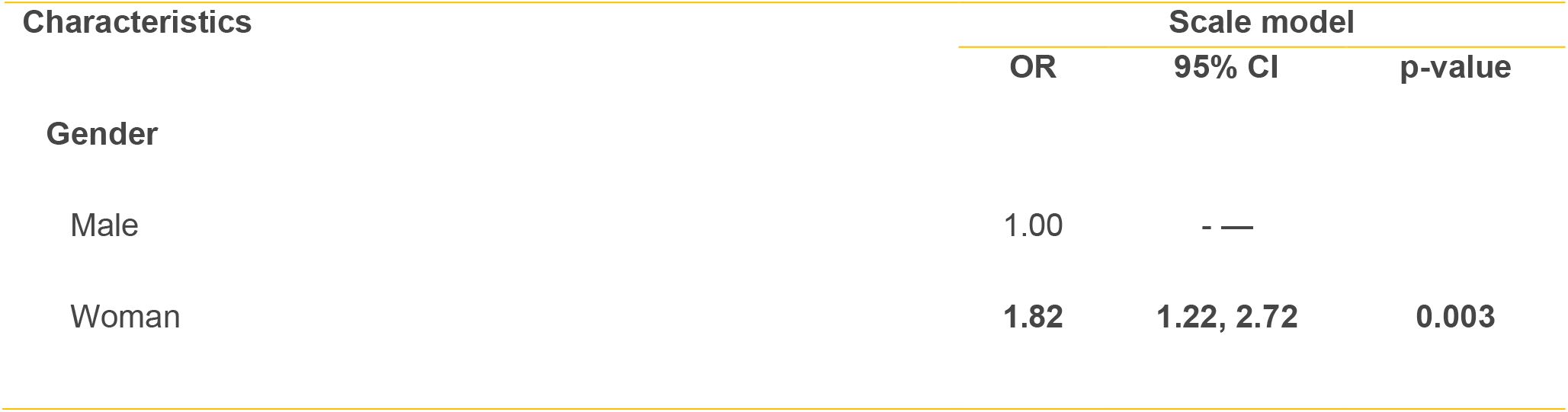
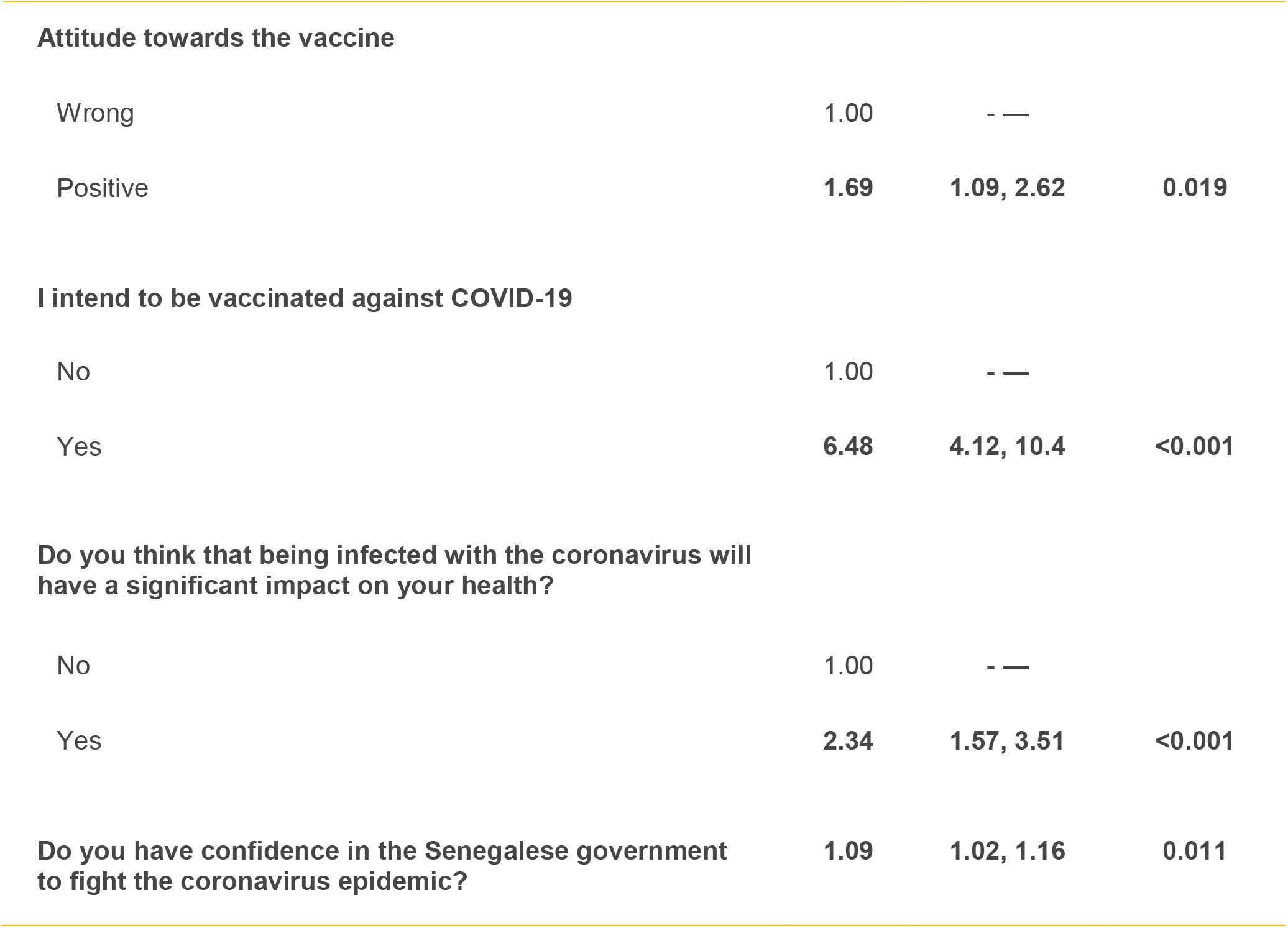
Results of the multivariate analysis

| Characteristics | Scale model |  |  |
| --- | --- | --- | --- |
|  | OR | 95% CI | p-value |
| **Gender** |  |  |  |
| Male | 1.00 | - — |  |
| Woman | 1.82 | 1.22, 2.72 | 0.003 |
| **Attitude towards the vaccine** |  |  |  |
| Wrong | 1.00 | - — |  |
| Positive | **1.69** | **1.09, 2.62** | **0.019** |
| **I intend to be vaccinated against COVID-19** |  |  |  |
| No | 1.00 | - — |  |
| Yes | **6.48** | **4.12, 10.4** | **<0.001** |
| **Do you think that being infected with the coronavirus will have a significant impact on your health?** |  |  |  |
| No | 1.00 | - — |  |
| Yes | **2.34** | **1.57, 3.51** | **<0.001** |
| **Do you have confidence in the Senegalese government to fight the coronavirus epidemic?** | **1.09** | **1.02, 1.16** | **0.011** |

## 4. Discussion

The results of this research are important in the context of the many controversies surrounding various vaccines, including AstraZeneca’s vaccine, which is at the heart of the COVAX initiative for Africa. As Senegal began administering the Sinopharm vaccine in early March 2021 and then its first doses of AstraZeneca, understanding people’s perceptions of vaccine research is essential. Indeed, in a context where there are calls for the decolonisation of global health research and for more vaccine research to be conducted in Africa [13,18], obtaining the views of those affected is an essential ethical issue [28,29].

In the contemporary context of this study, it is not surprising that the most favourable factor for wanting to participate in COVID-19 vaccine research is having the intention to be vaccinated. It should be noted that we conducted this study before the vaccines arrived in Senegal. The study also shows that other factors related to the health of individuals affect the intention to participate in research, such as perceived health status, belief in the consequences of vaccination on health, and positive attitude towards vaccination. Indeed, several authors have pointed out the importance of the health status and risk perception of the people concerned in the context of controlled human infection studies like experimental vaccines for COVID-19 [29].

It is not only individual factors that seem to influence the intention of study participants. The research confirms the importance of the notion of trust, which is an essential value for the effectiveness of health systems [30] and for the intention to adopt health-promoting behaviours. As with the acceptability of government measures to control COVID-19 [31] or the intention to be vaccinated against COVID-19 [32], trust is an important determinant of willingness to participate in research [20]. Research in the United States of America has shown how racialised history affects African Americans’ trust in government and influenza vaccination [33]. COVID-19 was a reminder that democracy, health, and pandemic are interrelated [34]. In Senegal, this trust seems to be multifaceted since it is demanded not only for the vaccine product but also for the health personnel and the government. Trust was already at the centre of the relationship between researchers and the population during the hepatitis B vaccine trial in Senegal in the 1970s and 1980s [3]. These results seem logical in a global context where *fake news* is widespread about vaccines, where Africa has been the scene of numerous medical experimentation abuses [1,2], and where the current mistrust of the Senegalese state apparatus is significant, including in the context of the COVID-19 pandemic [35]. It is essential to restore or increase this multifaceted trust to improve the willingness of Senegalese to participate in vaccine research.

## Data Availability

Data are available upon request to the authors.

## Ethical approval

The study was approved by the National Ethics Committee for Health Research in Senegal (SEN/20/23).

---

**Appendix 1:**
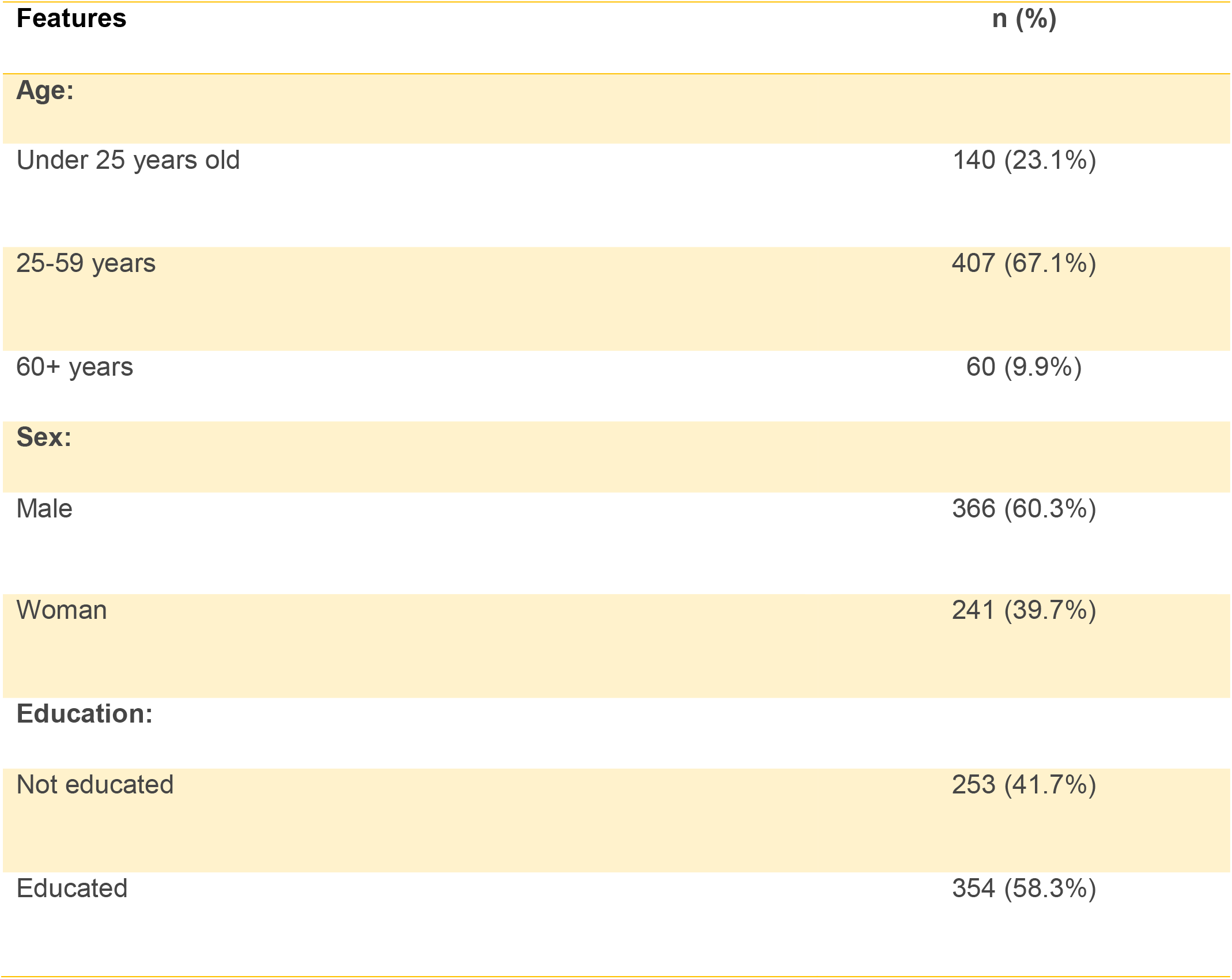
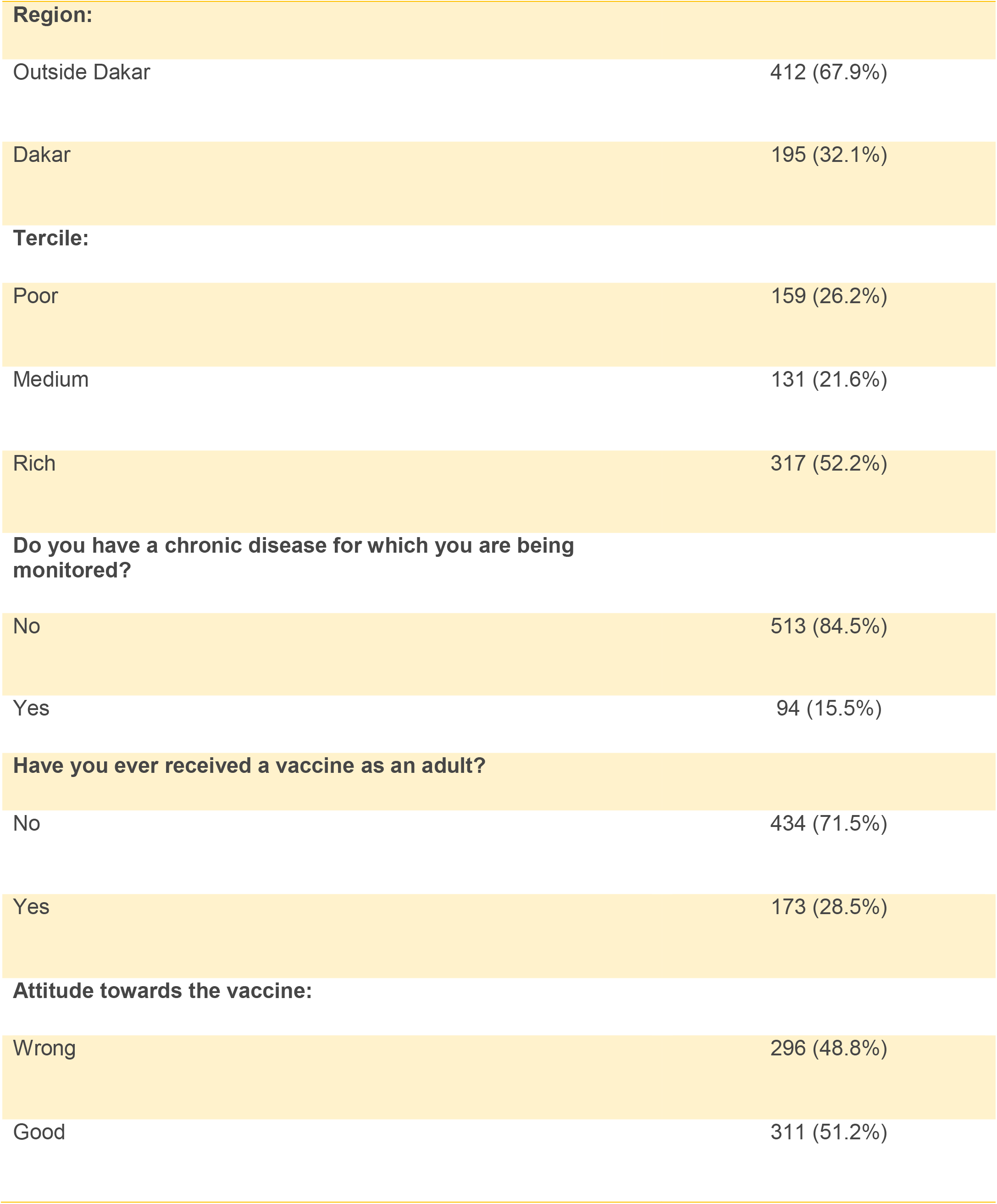
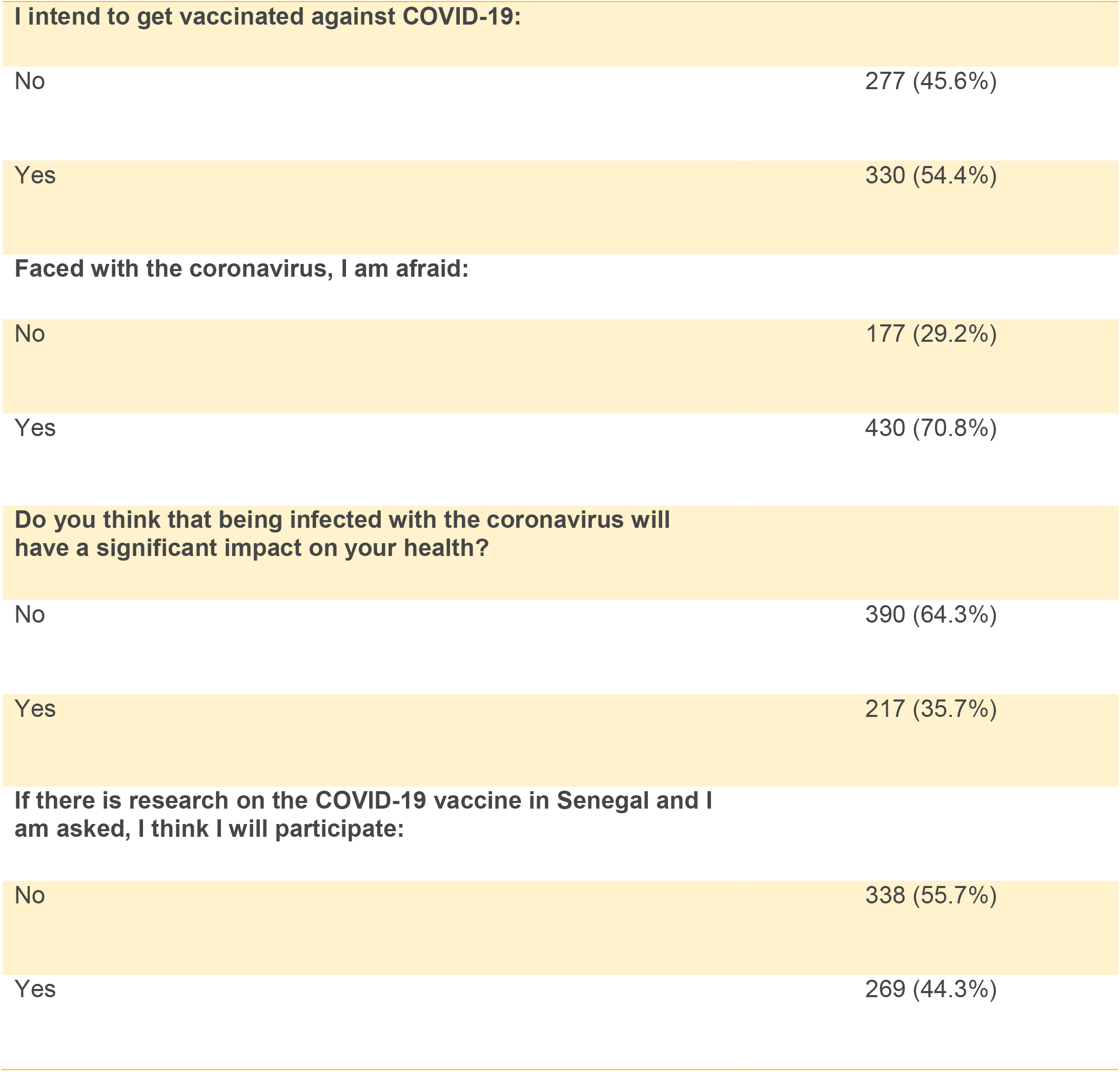
Distribution of respondents by characteristics (N=607)

**Appendix 2:**
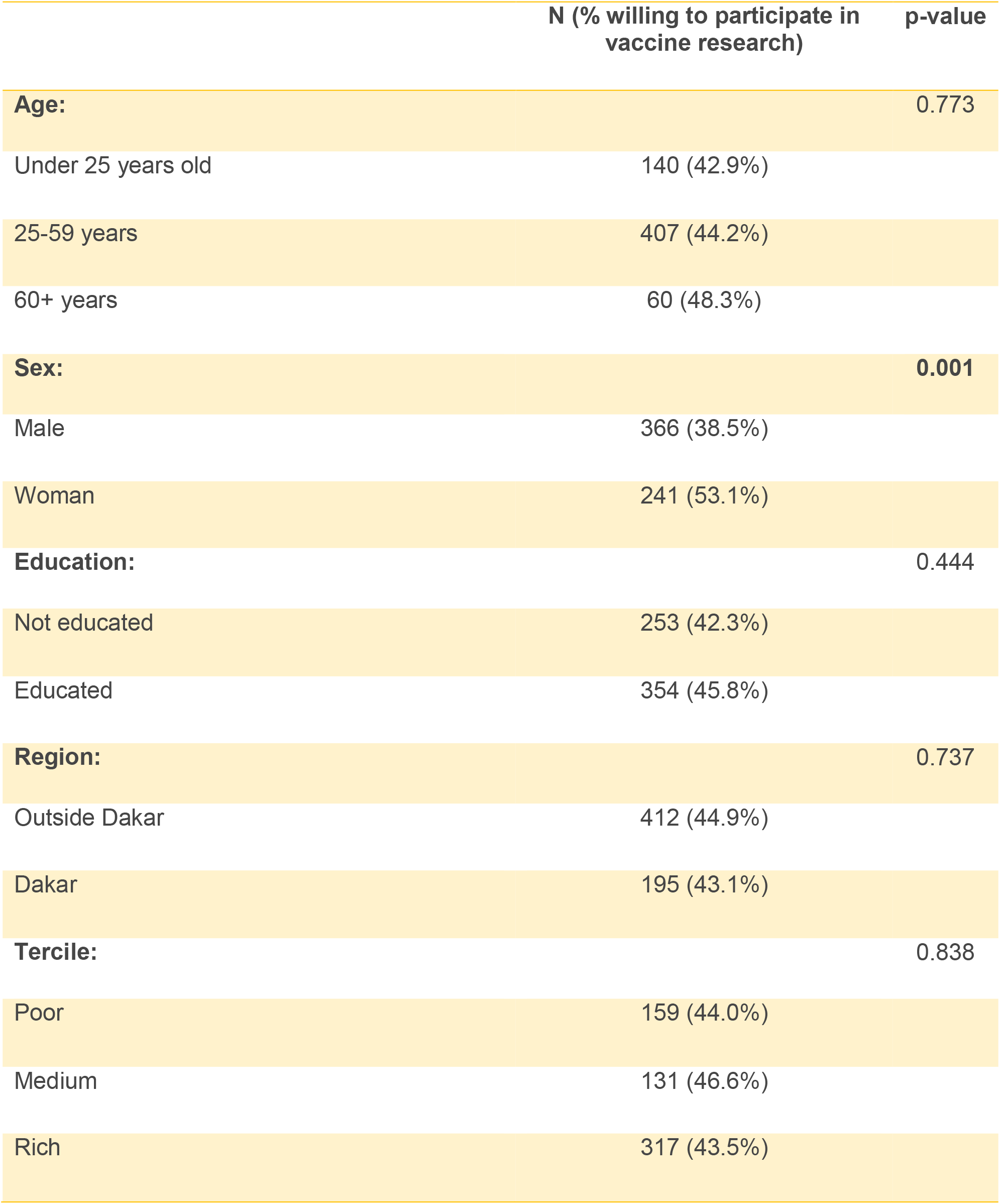
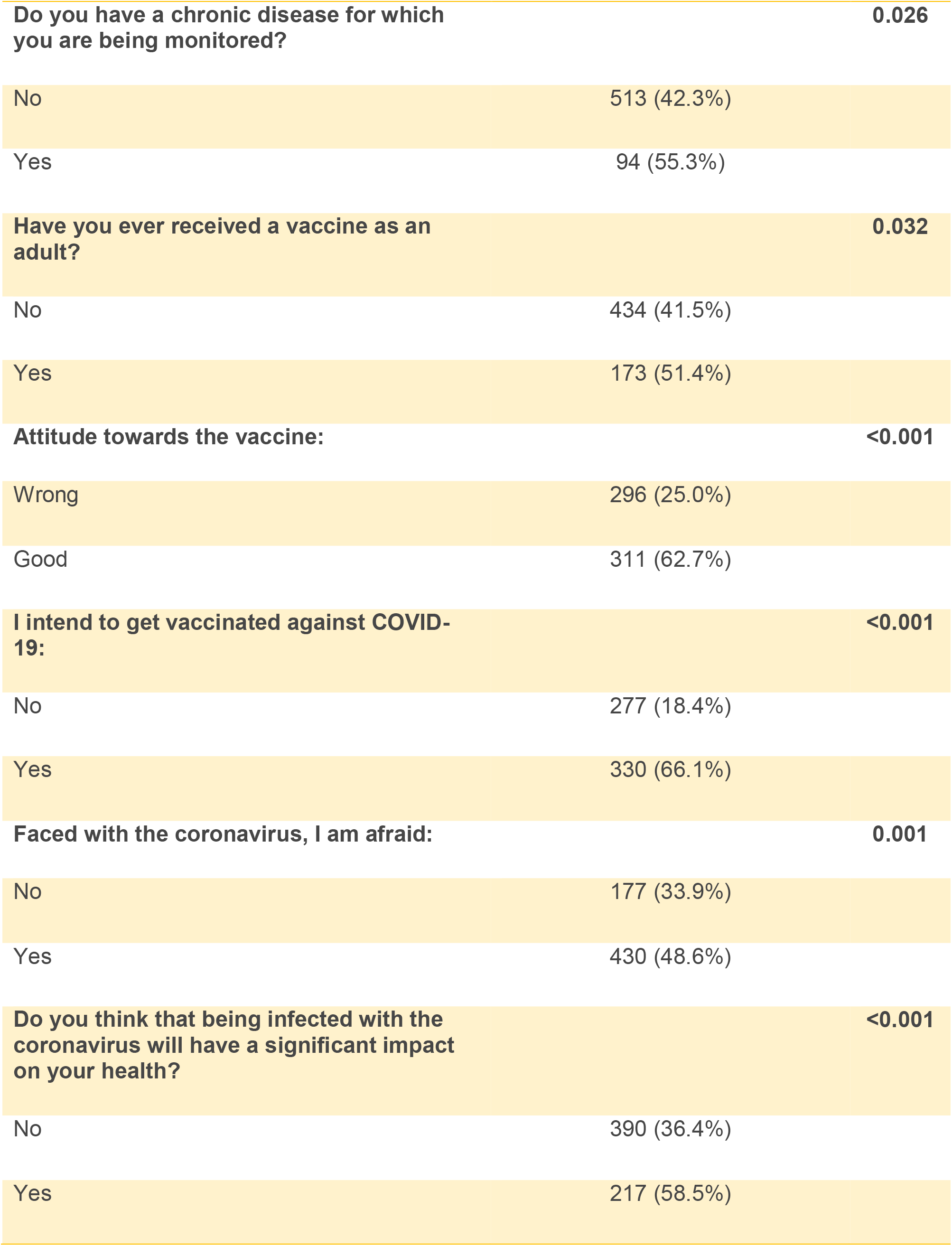
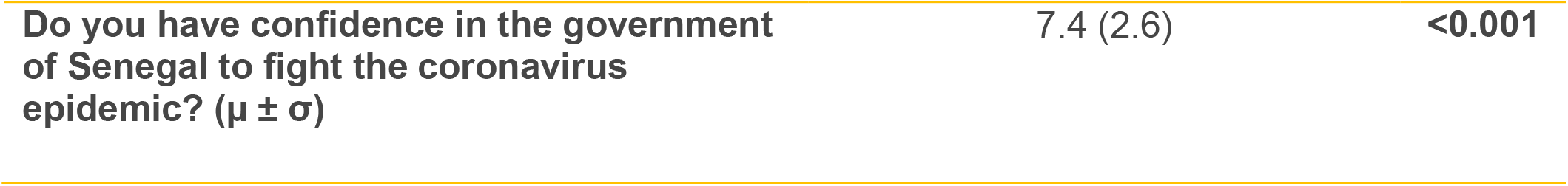
Distribution of respondents by characteristics and willingness to participate in COVID-19 vaccine research (N=607)

## Footnotes

1 https://covid19.trackvaccines.org/trials-vaccines-by-country/

